# Efficacy Validation of a Novel MRI-Based Whole-Body Rapid Bone Scan (WB-RBS) Strategy for Diagnosing Bone Metastases: A Prospective Trial

**DOI:** 10.64898/2026.05.17.26352855

**Authors:** Xusha Wu, Jian Zhang, Yifan He, Yang Zhang, Xiaowei Kang, Wenzhong Hu, Yan Li, Huiwen Ma, Yingxin Wang, Yu Song, Xiangxiang Chen, Fangjie Huo, Yi Zhang, Hong Yin, Yibin Xi

## Abstract

**Background:** Traditional bone scintigraphy for detecting malignant bone metastases is limited by suboptimal accuracy and radiation exposure. Whole-body magnetic resonance imaging (WB-MRI), while an alternative, requires lengthy scan times and high patient compliance.

**Purpose:** To develop a novel, rapid whole-body bone screening (WB-RBS) MRI protocol and evaluate its diagnostic performance for bone metastasis detection.

**Materials and Methods:** Patients with pathologically confirmed malignancies and healthy controls were prospectively enrolled. All participants underwent WB-RBS (acquisition time: about 10 min); patients additionally underwent WB-MRI (about 70 min). Three radiologists, blinded to clinical data, independently evaluated the images for bone metastases. A consensus expert diagnosis served as the reference standard to calculate the diagnostic performance of WB-RBS. Specificity was further assessed in the healthy control group.

**Results:** Seventy patients and 19 healthy controls were included. WB-RBS demonstrated excellent inter-reader agreement at the patient level. Compared with the reference standard, WB-RBS achieved an accuracy of 77.1%–91.4% at the patient level and a slightly lower accuracy (70.6%–82.5%) at the lesion level. At diagnostic confidence thresholds 1–3, the correlations between WB-RBS ratings and the reference standard were statistically significant for both patient- and lesion-level analyses.

**Conclusion:** WB-RBS showed favorable inter-reader agreement and high accuracy for bone metastasis screening at the patient level, while substantially reducing scan time and cost. Its rapid, radiation-free nature and high accessibility offer distinct clinical advantages, supporting its potential as an alternative screening tool to conventional bone scintigraphy.

**Summary:** For patients requiring assessment of bone metastases, the WB-RBS protocol provides a rapid (about 10-min), radiation-free, and accurate diagnostic option that facilitates clinical decision-making and enhances patient comfort.

**Key Points:**

- This study addresses the clinical need for a fast, accurate, and radiation-free bone metastasis screening tool by proposing a novel whole-body MRI-based screening strategy, which overcomes the limitations of conventional bone scintigraphy and reduces the lengthy acquisition time associated with standard WB-MRI.
- The approximately 10 minutes WB-RBS protocol developed demonstrated high diagnostic performance in detecting bone metastases in cancer patients, with favorable diagnostic accuracy.
- The protocol exhibits outstanding diagnostic capability for long bones of limbs and lesions larger than 1.5 cm in diameter, providing a practical and efficient imaging tool for identification and localization of bone metastases.

---

Bone is one of the most frequent sites of metastasis in cancer progression, significantly impacting patients’ quality of life and prognosis (1). In adults aged 25 years and older, lung cancer is the most common primary malignancy leading to bone metastases, followed by prostate and breast cancers (1). The cornerstone of managing metastatic bone disease lies in early and accurate screening, along with timely intervention, which is essential for preventing severe skeletal-related events (SREs) such as disabling pain, hypercalcemia, pathological fractures, or spinal cord compression (2).

However, current mainstream screening strategies for bone metastases face substantial health-economic and technical limitations. In modern clinical practice, technetium-99m-labeled bisphosphonate bone scintigraphy (BS) remains widely used as an initial screening tool due to its relative speed (3). Nevertheless, its diagnostic accuracy remains suboptimal and is highly susceptible to subjective interpretation, largely owing to its limited ability to provide precise anatomical localization of lesions (4, 5).

Whole-body magnetic resonance imaging (WB-MRI) has been established as a reference method with higher sensitivity, specificity, and the advantage of being radiation-free (5–17). However, its standard protocol is time-consuming, which can reduce patient compliance, introduce motion artifacts, and impose significant temporal and economic burdens, thereby limiting its widespread applicability in screening settings (7, 18). Consequently, there is a clear clinical need for a radiation-free screening tool that combines the accuracy of WB-MRI with the speed of BS.

Previous studies have primarily focused on accelerating scanning speed or optimizing sequence combinations. For instance, rapid T1 Dixon sequences have been employed to evaluate bone metastases in prostate cancer (19), while reduced WB-MRI protocol combinations have been validated in both prostate cancer and multiple myeloma (20). Early attempts also implemented echo-planar imaging to achieve whole-body rapid scanning. to evaluate bone metastases (21, 22). However, these approaches often require breath-holding and are susceptible to motion artifacts, are limited to specific diseases lacking generalizability, or significantly compromise spatial resolution and signal-to-noise ratio while introducing susceptibility artifacts. Consequently, they have failed to provide a fundamentally alternative bone-screening solution that balances accessibility, generalizability, and diagnostic accuracy in resource-constrained settings.

Therefore, this study aimed to develop and validate a novel, rapid whole-body bone screening (WB-RBS) MRI protocol designed to substantially improve diagnostic accuracy while maintaining screening efficiency. We employed a prospective design, using the diagnostically superior WB-MRI as the reference standard to evaluate the performance of the new protocol. If its diagnostic value is confirmed, this study will provide empirical support for WB-RBS as a superior skeletal screening tool capable of replacing conventional BS.

## Materials and Methods

### Study design

This prospective study enrolled patients between January 2025 and December 2025 who met the following criteria: age ≥18 years, histopathologically confirmed malignancy with a high risk of bone metastasis, and completion of both non-contrast WB-MRI and the developed MRI-based WB-RBS examination. Patients unable to maintain a supine position or those with metallic implants were excluded. Additionally, healthy controls with no history of malignancy or other bone-affecting diseases were recruited. A participant screening flowchart is presented in Figure 1.

**Figure 1.**
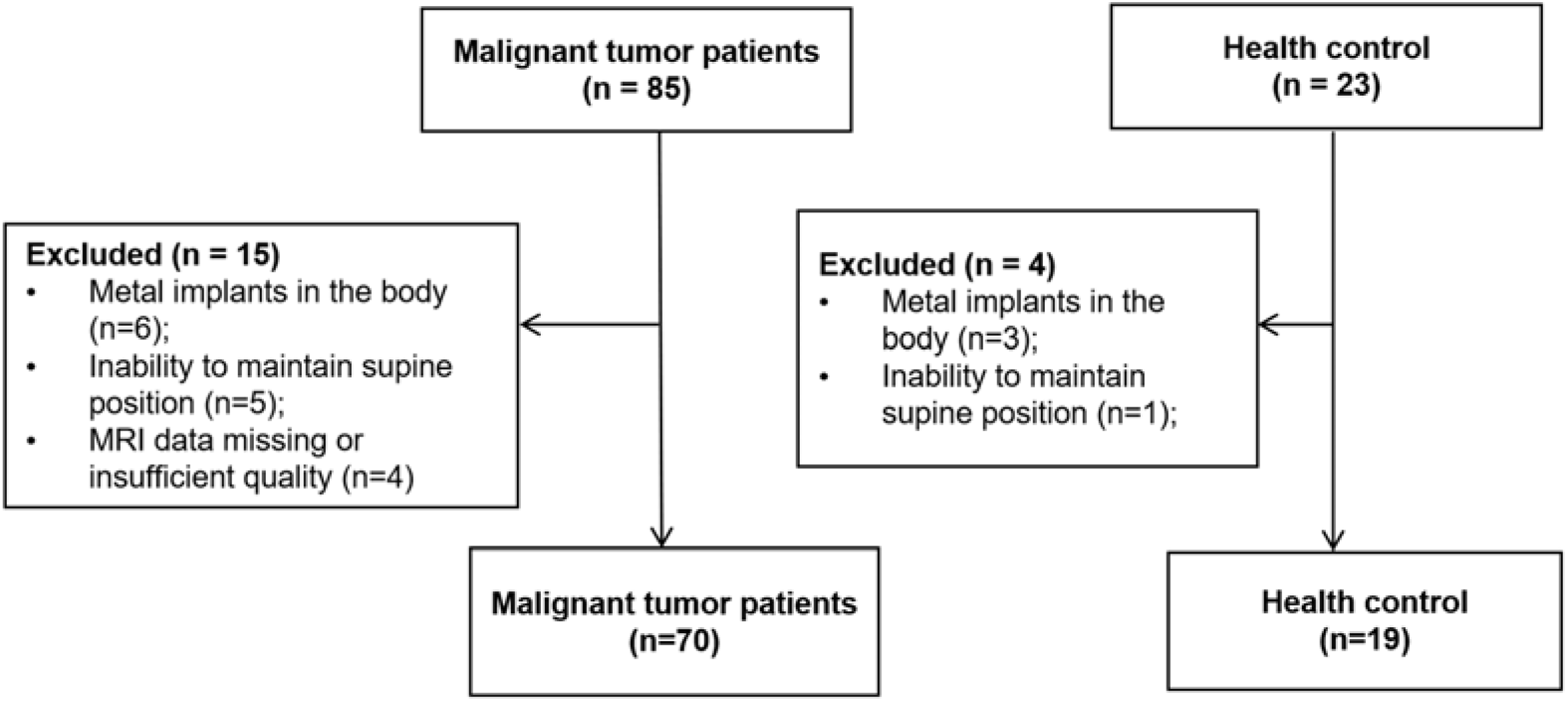
Flowchart illustrating the enrollment of malignant tumor patients and healthy controls.

The study was conducted in accordance with the ethical principles of the Declaration of Helsinki and all applicable local regulations. The study protocol was approved by the Institutional Review Board, and written informed consent was obtained from all participants.

### Data Collection

All participants underwent WB-RBS. Additionally, the patient cohort completed WB-MRI examinations. The same combined WB-RBS and WB-MRI protocols were performed during patient follow-up visits. Baseline clinical information, including age, sex, and primary cancer type, was obtained from medical records for participant characterization. Imaging was performed using a Siemens 3.0 T Prisma scanner equipped with a 20-channel head-neck coil, two 18-channel body coils, and a 32-channel spine coil. The WB-RBS protocol (approximately 10 minutes) and WB-MRI protocol (approximately 70 minutes) are detailed in the Supplementary Material and Supplementary Table S1.

### Lesion Assessment

All patients were randomly coded and anonymized. Three radiologists with different seniority (designated as A, B, and C, corresponding to name initials Y.X., X.K., and Y.Z., with 20, 12, and 9 years of diagnostic experience, respectively) independently evaluated metastases on WB-RBS and WB-MRI images, blinded to clinical data of the patients. The assessment covered the skull, spine, thorax and shoulder girdle, pelvis, and long bones of the limbs (including the radius/ulna and tibia/fibula). Evaluation began with the WB-RBS images. After a 3-week washout period, the radiologists then assessed the WB-MRI images, which were presented in a newly randomized order.

All WB-RBS images were evaluated by the same three radiologists, who had undergone standardized training, using a consensus reading method via a PACS system. The assessment process consisted of two steps. First, potential lesions were identified based on consensus-defined imaging features (e.g., round or patchy high signal on WB-RBS with ill-defined margins, cortical disruption, or focal bone defect; see Figure 2). Second, each identified lesion was assigned a subjective probability score for metastasis (1: >90%, definite; 2: 70–90%, probable; 3: 50–70%, equivocal; 4: ≤50%, unlikely). To investigate diagnostic performance across confidence levels, four predefined binary classification thresholds (Supplementary Table S2) were applied to convert these probability scores into final “metastatic” or “non-metastatic” diagnoses.

**Figure 2.**
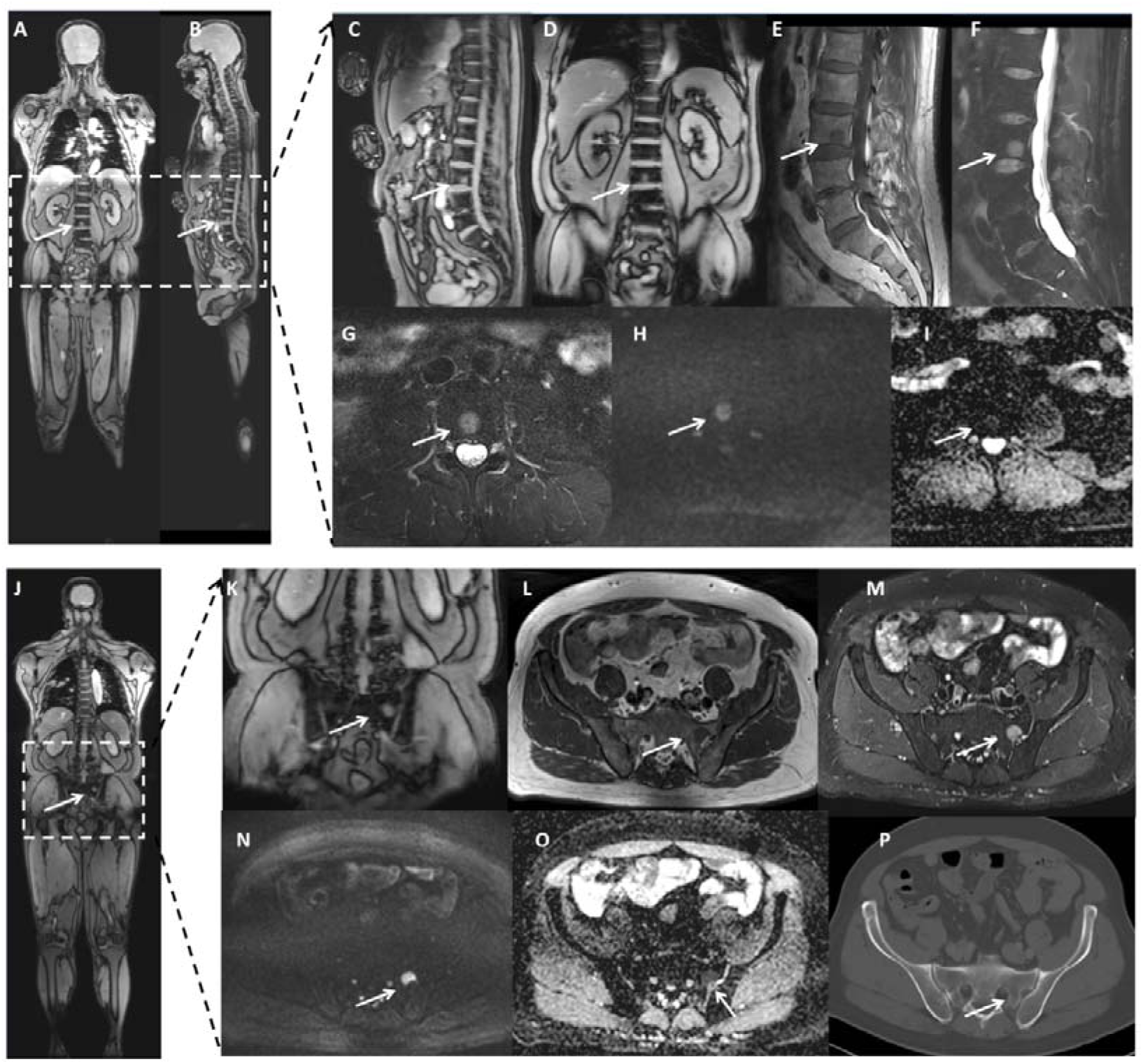
An elderly male patient diagnosed with small cell lung cancer one week prior. (A-D) Whole-body rapid bone scan (WB-RBS) sagittal and coronal whole body and partial views demonstrate an ovoid, hyperintense focus within the lumbar vertebrae 3 vertebral body (score 1 point). (E-G) Sagittal and axial lumbar spine MRI sequences reveal an ovoid lesion exhibiting T1 hypointensity and fat-suppressed T2 hyperintensity at lumbar vertebrae 3. (H, I) Corresponding diffusion-weighted imaging (DWI) with high b-value shows hyperintensity, while the apparent diffusion coefficient (ADC) map exhibits hypointensity, indicating restricted diffusion and suggesting a metastatic bone lesion. (J, K) WB-RBS coronal whole body and partial view demonstrates an ovoid, hyperintense focus on the left side of the sacrum (score 1 point). (L, M) Axial pelvic MRI sequences reveal an ovoid lesion exhibiting T1 hypointensity and fat-suppressed T2 hyperintensity on the left side of the sacrum. (N, O) Corresponding DWI with high b-value shows hyperintensity and the ADC map shows hypointensity, indicating restricted diffusion and suggesting a metastatic bone lesion. (P) Corresponding pelvic CT image shows no definite evidence of bone destruction.

Cases with more than 5 metastatic foci in a single anatomical region (e.g., entire spine) or a total of more than 20 identifiable metastatic lesions throughout the body were defined as having diffuse metastatic disease. For these cases, a sampling rule was applied: within each predefined anatomical region (skull, spine, thorax/shoulder girdle, pelvis, long bones of limbs), the three largest lesions (by maximum diameter) were recorded and included in the analysis. If a region contained fewer than 3 lesions, all were recorded. Additional smaller, diffusely distributed lesions within the same region were documented as “diffuse involvement” but were not individually measured or counted.

This study aimed to evaluate the diagnostic performance of WB-RBS for detecting hematogenously disseminated bone metastases. Therefore, cases of bone destruction due to direct tumor invasion were explicitly excluded from the analysis.

### Defining Reference Standard

The reference standard for bone metastasis—the final diagnosis (metastatic or non-metastatic) for all patients—was established through a consensus discussion among the three radiologists after completion of the WB-RBS and WB-MRI lesion assessments. This standard was primarily based on WB-MRI findings. In cases of atypical or equivocal WB-MRI presentations, additional available imaging modalities (such as CT, PET/CT, or BS) and follow-up imaging data for the same patient were integrated to confirm the final diagnosis, ensuring the reliability of the reference standard. This process yielded a definitive metastatic or non-metastatic classification for each lesion, which was subsequently used to evaluate the diagnostic performance of WB-RBS.

### Stratified Analyses

To systematically evaluate the impact of lesion size on the diagnostic performance of WB-RBS, lesions were stratified into two groups based on the maximum diameter measured on the reference standard WB-MRI: small lesions (≤1.5 cm) and large lesions (>1.5 cm). This stratified design allowed for an explicit examination of the relationship between WB-RBS performance and lesion size, quantifying its detection capability for lesions of different dimensions encountered in clinical practice.

Furthermore, lesions were categorized into the following five anatomical groups for evaluation: skull, thorax and shoulder girdle (sternum, ribs, clavicles, and scapulae), spine, pelvis, and long bones of the limbs (including humerus, radius/ulna, femur, and tibia/fibula). This stratified design enabled a precise analysis of the diagnostic performance of WB-RBS across different anatomical regions, clarifying its strengths and limitations in screening for metastases at various sites.

### Statistical Analysis

Statistical analyses were performed using SPSS software (version 25.0; IBM Corp.), with a two-sided *p* value < 0.05 considered statistically significant. Descriptive statistics summarized clinical and imaging characteristics, presented as mean ± standard deviation (SD), range, or percentage. Inter-reader agreement among the three readers for WB-RBS assessments was evaluated using Fleiss’ kappa at both the patient level (presence/absence of metastasis per patient) and the lesion level (assessment per lesion). The agreement between the WB-RBS and WB-MRI protocols was assessed using Cohen’s kappa (interpretation: <0.20, poor; 0.21–0.40, fair; 0.41–0.60, moderate; 0.61–0.80, substantial; >0.80, almost perfect). Diagnostic performance metrics (sensitivity, specificity, positive predictive value, negative predictive value, and accuracy) for WB-RBS were calculated against the reference standard at both the patient and lesion levels. Specificity and negative predictive value were also calculated within the healthy control group. Further stratified evaluations were performed based on lesion size and anatomical location. Additionally, the discriminatory ability of the three readers across the four scoring thresholds was analyzed using ROC curves, and the area under the curve (AUC) was calculated. Spearman’s rank correlation analysis was used to explore the correlation between WB-RBS results and the reference standard at the different thresholds.

## Results

### Demographic and clinical data

Between January 2025 and December 2025, 85 patients with malignancies were enrolled. Fifteen patients were excluded for the following reasons: presence of metallic implants (n=6), inability to maintain a supine position (n=5), and missing or poor-quality MRI data (n=4) (Figure 1). All malignant diagnoses were histopathologically confirmed via biopsy or surgical resection. The final cohort comprised 70 patients (mean age 64.1 ± 10.7 years; 47 men, 23 women), including 65 with lung cancer, 2 with esophageal cancer, 2 with breast cancer, and 1 with thymic carcinoma. During the same period, 23 healthy volunteers were recruited, with 19 completing the study (mean age 58.6 ± 10.0 years; 7 men, 12 women). Participant demographics and clinical characteristics are detailed in Table 1. Details regarding baseline and follow-up MRI scans are provided in Supplementary Table S3.

**Table 1.**
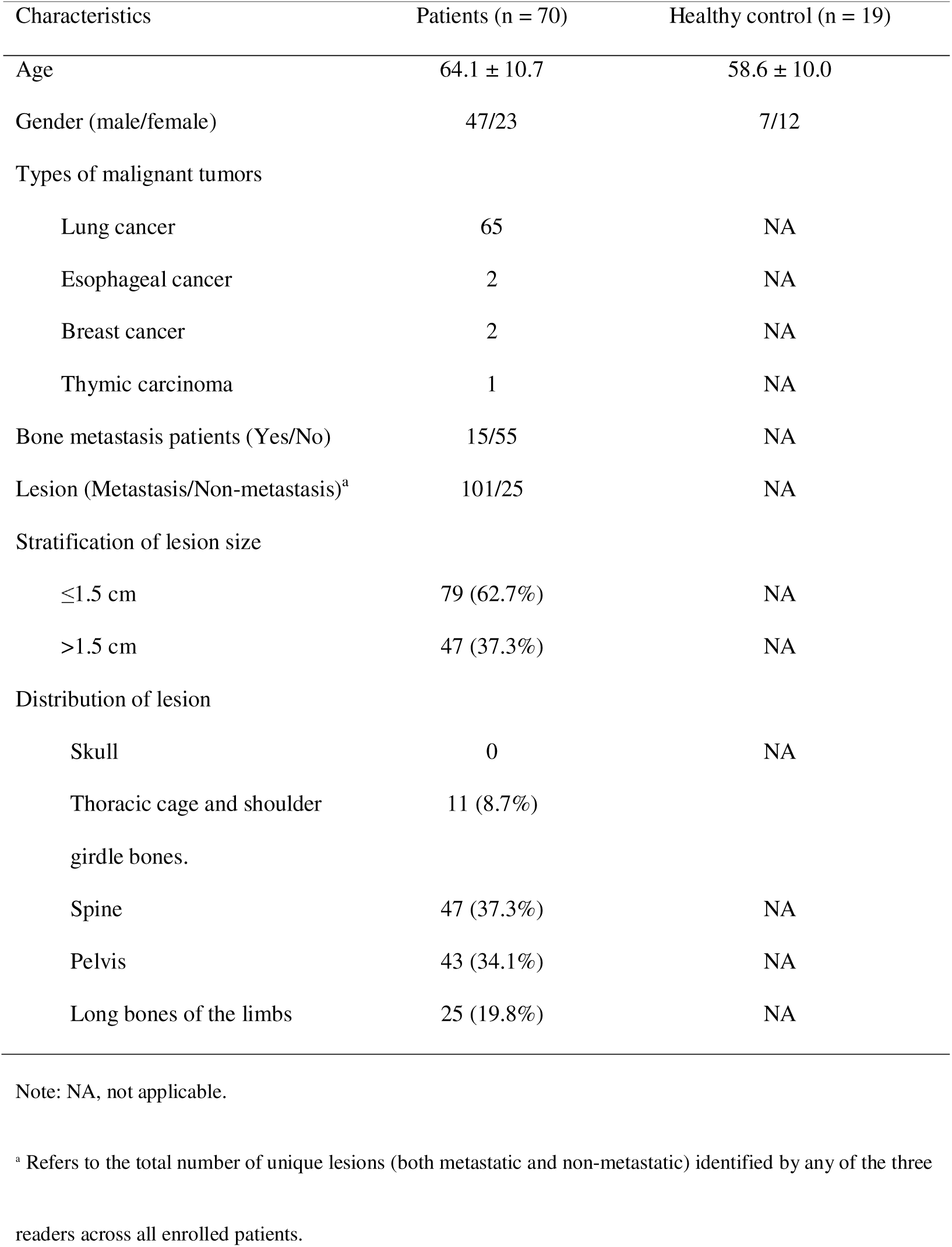
Demographic and clinical characteristics of participants.

| Characteristics | Patients (n = 70) | Healthy control (n = 19) |
| --- | --- | --- |
| Age | 64.1 ± 10.7 | 58.6 ± 10.0 |
| Gender (male/female) | 47/23 | 7/12 |
| Types of malignant tumors |  |  |
| Lung cancer | 65 | NA |
| Esophageal cancer | 2 | NA |
| Breast cancer | 2 | NA |
| Thymic carcinoma | 1 | NA |
| Bone metastasis patients (Yes/No) | 15/55 | NA |
| Lesion (Metastasis/Non-metastasis)<sup>a</sup> | 101/25 | NA |
| Stratification of lesion size |  |  |
| ≤1.5 cm | 79 (62.7%) | NA |
| >1.5 cm | 47 (37.3%) | NA |
| Distribution of lesion |  |  |
| Skull | 0 | NA |
| Thoracic cage and shoulder<br>girdle bones. | 11 (8.7%) |  |
| Spine | 47 (37.3%) | NA |
| Pelvis | 43 (34.1%) | NA |
| Long bones of the limbs | 25 (19.8%) | NA |

Note: NA, not applicable.
<sup>a</sup> Refers to the total number of unique lesions (both metastatic and non-metastatic) identified by any of the three
readers across all enrolled patients.

Following the consensus review, 15 patients were confirmed to have bone metastases, among whom 6 were classified as having diffuse metastatic disease, while 55 patients were metastasis-free. A total of 126 lesions were identified across all readers, including 101 metastatic and 25 non-metastatic lesions. The number of lesions detected by the three radiologists under different thresholds for both the WB-MRI and WB-RBS protocols are presented in Table 2. The distribution of lesions by size and anatomical location is summarized in Table 1.

**Table 2.** Number of lesions detected by three readers with two protocols and four thresholds.

| Threshold | Protocol | Reader A | Reader B | Reader C |
| --- | --- | --- | --- | --- |
| Thr 1 | WB-RBS | 86 | 83 | 86 |
|  | WB-MRI | 99 | 97 | 97 |
| Thr 2 | WB-RBS | 9 | 13 | 7 |
|  | WB-MRI | 1 | 3 | 3 |
| Thr 3 | WB-RBS | 6 | 8 | 8 |
|  | WB-MRI | 1 | 0 | 1 |
| Thr 4 | WB-RBS | 5 | 6 | 5 |
|  | WB-MRI | 0 | 2 | 2 |

Note: Thr, threshold.

### Agreement Analysis

Inter-reader agreement for the WB-RBS protocol, assessed at both the patient and lesion levels, demonstrated excellent consistency at the patient level (Figure 3, Supplementary Table S4). Fleiss’ kappa values for WB-RBS at the patient level were ≥0.86 across all confidence thresholds (Thresholds 1-4). At the lesion level, substantial agreement was maintained, with Fleiss’ kappa values ranging from 0.77 to 0.84. The three readers also showed high inter-reader agreement in their assessments of WB-MRI at both levels.

**Figure 3.**
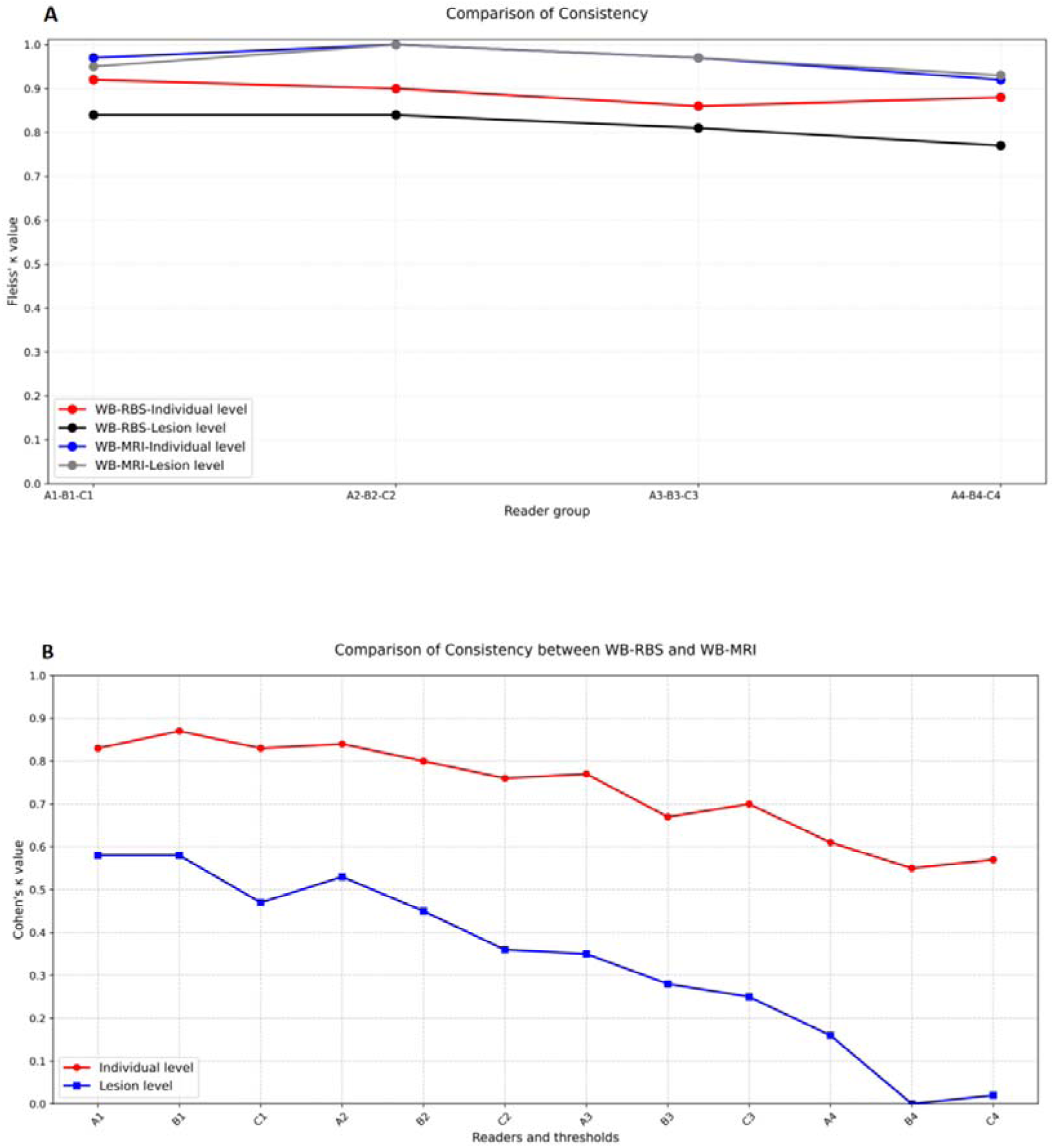
(A) Line graphs depicting Kappa values for inter-reader agreement among three readers on per-patient and per-lesion assessments using the WB-MRI and WB-RBS protocols at different thresholds. (B) Line graphs depicting Kappa values for agreement between the WB-RBS and WB-MRI protocols on per-patient and per-lesion assessments by three readers at different thresholds. A1-4 indicates the results of Reader A using thresholds 1-4, and B1-4 and C1-4 are similar.

Analysis of agreement between WB-RBS and WB-MRI for each reader showed that at the patient level, agreement was highest at Threshold 1 and decreased with increasing threshold stringency (Kappa range: 0.55-0.87, all p < 0.05) (Figure 3, Supplementary Table S5). A similar trend was observed at the lesion level, where agreement declined with higher thresholds. Kappa values ranged from 0.25 to 0.58 (all p < 0.05) for Thresholds 1-3, but were non-significant at Threshold 4 (Kappa range: 0.00-0.16, all p > 0.05).

### The Diagnostic Performance of the WB-RBS Protocol

The diagnostic performance of the WB-RBS protocol, as evaluated by the three readers across different thresholds at the patient and lesion levels, is presented in Table 3 and Figure 4. At the patient level, comparison with the reference standard revealed specificities ≥78.2% for all readers across thresholds. Patient-level accuracy ranged from 77.1% to 91.4%, decreasing with higher thresholds. At Threshold 1, all three readers achieved accuracy ≥90%. At the lesion level, specificity was ≥77.2%, while sensitivity decreased with higher thresholds. Lesion-level accuracy ranged from 70.6% to 82.5%, peaking for all readers at Threshold 2. The diagnostic performance of WB-MRI is also provided for reference (Supplementary Table S4). Specificity of WB-RBS in healthy individuals is presented in the supplementary information.

**Figure 4.**
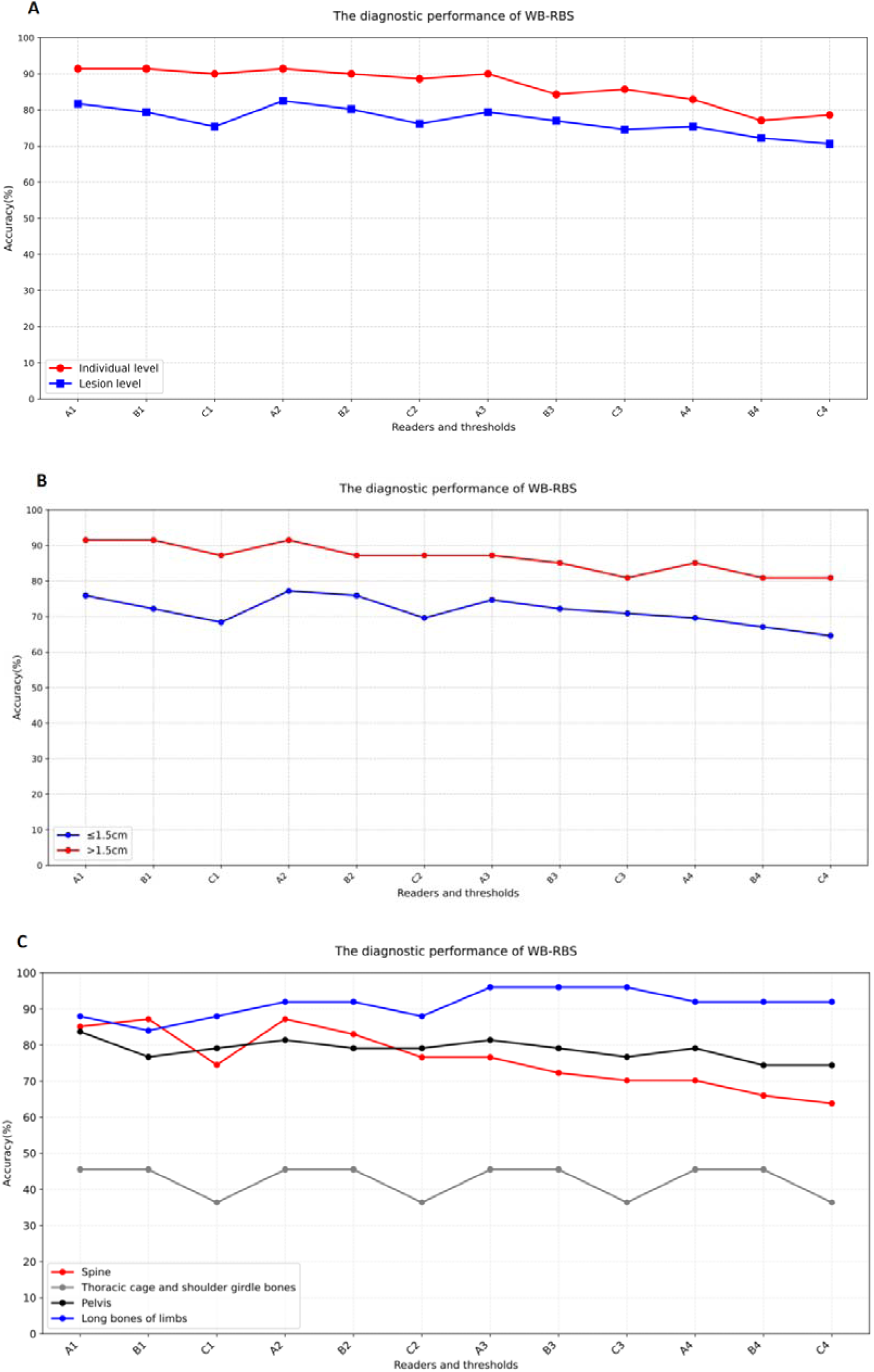
(A) Line graphs depicting the diagnostic accuracy of the WB-RBS protocol by three readers on both per-patient and per-lesion bases at different thresholds. (B) Line graphs depicting the diagnostic accuracy of the WB-RBS protocol by three readers for lesions of different sizes at different thresholds. (C) Line graphs depicting the diagnostic accuracy of the WB-RBS protocol by three readers for lesions in different locations at different thresholds. A1-4 indicates the results of Reader A using thresholds 1-4, and B1-4 and C1-4 are similar.

**Table 3.** Diagnostic performance of the WB-RBS protocol evaluated by three readers at different thresholds based on the patient level and lesion level.

| Readers | Level | Sensitivity (%) | Specificity (%) | PPV (%) | NPV (%) | Accuracy (%) |
| --- | --- | --- | --- | --- | --- | --- |
| A1 | Patient level | 80.0 | 94.5 | 80.0 | 94.5 | 91.4 |
| A2 | Patient level | 86.7 | 92.7 | 76.5 | 96.2 | 91.4 |
| A3 | Patient level | 86.7 | 90.9 | 72.2 | 96.2 | 90.0 |
| A4 | Patient level | 86.7 | 81.8 | 56.5 | 95.7 | 82.9 |
| B1 | Patient level | 80.0 | 94.5 | 80.0 | 94.5 | 91.4 |
| B2 | Patient level | 86.7 | 90.9 | 72.2 | 96.2 | 90.0 |
| B3 | Patient level | 86.7 | 83.6 | 59.1 | 95.8 | 84.3 |
| B4 | Patient level | 86.7 | 74.5 | 48.1 | 95.3 | 77.1 |
| C1 | Patient level | 80.0 | 92.7 | 75.0 | 94.4 | 90.0 |
| C2 | Patient level | 80.0 | 90.9 | 70.6 | 94.3 | 88.6 |
| C3 | Patient level | 86.7 | 85.5 | 61.9 | 95.9 | 85.7 |
| C4 | Patient level | 86.7 | 76.4 | 50.0 | 95.5 | 78.6 |
| A1 | Lesion level | 81.2 | 84.0 | 95.3 | 52.5 | 81.7 |
| A2 | Lesion level | 86.1 | 68.0 | 91.6 | 54.8 | 82.5 |
| A3 | Lesion level | 87.1 | 48.0 | 87.1 | 48.0 | 79.4 |
| A4 | Lesion level | 87.1 | 28.0 | 83.0 | 35.0 | 75.4 |
| B1 | Lesion level | 78.2 | 84.0 | 95.2 | 48.8 | 79.4 |
| B2 | Lesion level | 85.1 | 60.0 | 89.6 | 50.0 | 80.2 |
| B3 | Lesion level | 87.1 | 36.0 | 84.6 | 40.9 | 77.0 |
| B4 | Lesion level | 87.1 | 12.0 | 80.0 | 18.8 | 72.2 |
| C1 | Lesion level | 77.2 | 68.0 | 90.7 | 42.5 | 75.4 |
| C2 | Lesion level | 81.2 | 56.0 | 88.2 | 42.4 | 76.2 |
| C3 | Lesion level | 84.2 | 36.0 | 84.2 | 36.0 | 74.6 |
| C4 | Lesion level | 84.2 | 16.0 | 80.2 | 20.0 | 70.6 |

Note: PPV, positive predictive value; NPV, negative predictive value.

### Diagnostic Performance of WB-RBS Based on Lesion Size and Location

All identified lesions were measured on WB-MRI and stratified by size: 79 lesions (62.7%) were small (≤1.5 cm), including 31 sub-centimeter (<1 cm) lesions, and 47 lesions (37.4%) were large (>1.5 cm). For large lesions, all readers achieved sensitivities of 95% across all thresholds, with accuracies ranging from 80.9% to 91.5%. For small lesions (≤1.5 cm), accuracies ranged from 64.6% to 77.2% across thresholds. Readers A and B achieved their highest accuracy at Threshold 2, while Reader C achieved it at Threshold 3. All readers demonstrated their lowest accuracy for small lesions at Threshold 4 (Figure 3, Supplementary Table S7).

The location-based stratified analysis showed that for metastases in the long bones of the limbs, specificities across the four thresholds ranged from 83.3% to 95.8%, with corresponding accuracies of 84%-96%. Diagnostic performance was moderate for spinal and pelvic lesions. Accuracy for spinal lesions tended to decrease with higher thresholds, while it remained relatively stable for pelvic lesions across readers and thresholds. The diagnostic performance of WB-RBS was lowest for lesions in the thoracic cage and shoulder girdle, with accuracies between 36.4% and 45.5%. No metastatic lesions were found in the skull. Detailed results are shown in Figure 4 and Supplementary Table S8.

### ROC Analysis

As shown in Figure 5, at the patient level, all three readers achieved AUCs above 0.85 for Thresholds 1-3. At the lesion level, Readers A and B maintained AUCs above 0.80 at Threshold 1, while Reader C’s performance was lower. Diagnostic accuracy was lowest at Threshold 4 for all readers at both patient and lesion levels.

**Figure 5.**
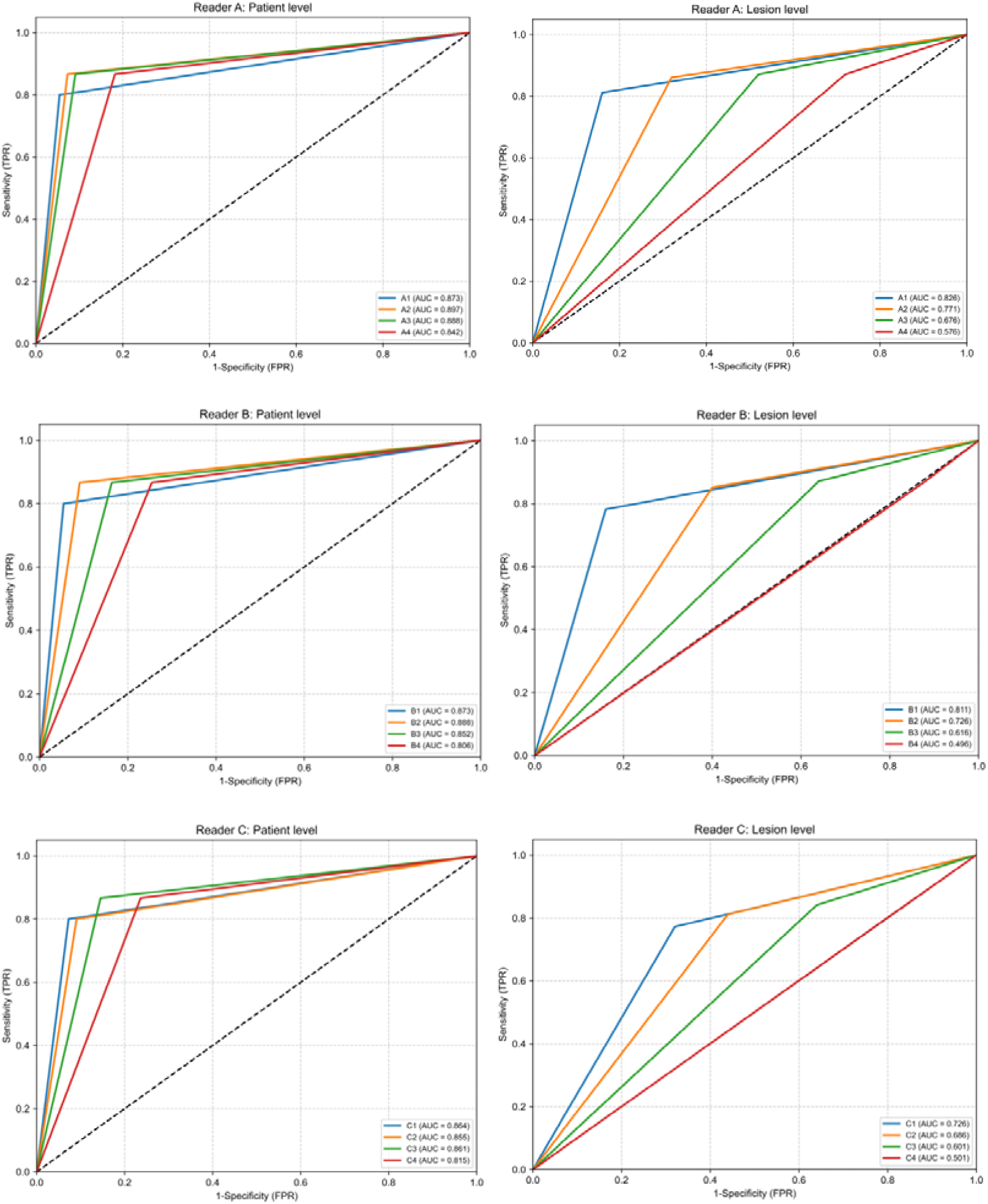
Receiver Operating Characteristic (ROC) curves of three readers at four thresholds, analyzed at both patient-level and lesion-level.

### Correlation Analysis

At the patient level, the diagnostic scores from all three readers showed statistically significant positive correlations with the reference standard across all four thresholds. At the lesion level, significant positive correlations were observed for Thresholds 1-3, while correlations were non-significant at Threshold 4 (Supplementary Table S9).

## Discussion

This prospective study demonstrates the significant advantages of a rapid MRI gradient-recalled-echo-based imaging technique for diagnosing bone metastases. The core innovation of WB-RBS lies in repurposing and optimizing a standard GRE sequence, originally intended for MRI localization, into a diagnostic sequence. This sequence offers fast imaging, free breathing, wide coverage, excellent compatibility, and low sensitivity to respiratory and cardiac motion artifacts, enabling a comprehensive, multi-planar skeletal evaluation. Compared to WB-MRI, the scan time was reduced by 85.7%. This innovative design facilitates the transition of MRI from a time-intensive advanced examination to an efficient screening tool, offering a novel technical pathway for the early diagnosis of bone metastases. Our prospective validation of WB-RBS for bone metastasis detection, through agreement and diagnostic performance analyses, demonstrates that the protocol exhibits good inter-reader agreement and robust diagnostic performance, particularly at the patient level, across confidence thresholds 1-3. The protocol is characterized by rapid scanning (approximately 10 minutes for full skeletal coverage), low cost, free breathing, and a radiation-free operation. Our findings may provide an effective alternative to current bone metastasis screening paradigms, potentially reshaping the screening workflow.

While previous studies have validated the diagnostic performance of WB-MRI (23–26), its implementation as a large-scale screening tool must consider cost-effectiveness (27), test efficiency, practical feasibility, impact on patient behavior, and integration into the overall clinical decision-making pathway (28). This study addresses a widespread clinical need: providing a convenient, dedicated bone-screening tool for patients with suspected bone metastases in settings where comprehensive WB-MRI is not routinely feasible. In contrast to previous studies that have primarily focused on increasing scanning speed or optimizing sequence combinations (19–22), WB-RBS is positioned as a targeted upgrade to conventional BS. Leveraging widely available MRI hardware and a brief acquisition time, it maintains critical diagnostic information and is particularly suited for resource-limited environments, scenarios requiring rapid screening, or patients intolerant of lengthy examinations, offering a pragmatic and potentially valuable tool for implementing precise bone metastasis assessment in real-world clinical practice.

In the agreement analysis, at the lesion level, when the most lenient diagnostic threshold (Threshold 4) was applied—classifying all indeterminate findings as positive—the interpretation variability among readers regarding the “uncertainty” boundary was maximized. Consequently, the agreement between WB-RBS and the reference standard was not statistically significant (p > 0.05). This reflects the fundamentally different objectives at this threshold: Threshold 4 aims for screening-level high sensitivity, causing the positive call rate of WB-RBS to be significantly higher than that of WB-MRI, which emphasizes specificity, thus systematically lowering agreement metrics. Notably, when more stringent diagnostic thresholds (Thresholds 1-3) were applied, the agreement became statistically significant and showed a progressively strengthening trend (all p < 0.05). This dynamic pattern demonstrates that WB-RBS can strike a balance between “high-sensitivity screening” and “high-certainty diagnosis” through threshold adjustment.

The stratified analyses revealed that WB-RBS performed best for large lesions and metastases in the long bones of the limbs, while its detection capability was weaker for small lesions and metastases in the thoracic cage and shoulder girdle. Sensitivity for small lesions ranged from 65.6% to 82%, whereas it reached 95% for the large-lesion group. Similarly, a previous large multicenter study reported a sensitivity of 82% for WB-MRI in patients with metastases ≥1 cm, but only 9% for smaller metastases (23). Furthermore, the WB-RBS protocol is particularly suitable for anatomically simpler regions. For instance, due to their clear boundaries and regular morphology, metastases in the long bones achieved accuracies of 84%-96% across all thresholds, suggesting that long-bone metastasis diagnosis may be the most reliable aspect of the WB-RBS protocol. The lowest diagnostic performance was observed for the thoracic cage and shoulder girdle (accuracy range: 36.4%-45.5%), which may be attributed to rib anatomy; a previous study on whole-body DWI reported a detection rate of only 11.9% for rib metastases (29). Future developments could focus on creating a dedicated AI-assisted bone metastasis detection module based on WB-RBS, utilizing deep learning to address the recognition challenges in the rib region. Standard WB-MRI protocols often fail to fully cover long bones, especially the upper limbs, potentially leading to missed metastases, while dedicated coverage increases scan time. In contrast, our WB-RBS protocol achieves truly comprehensive “whole-body” scanning, including the radius/ulna and tibia/fibula (as shown in the Supplementary Video S1 and S2). Therefore, the clinical utility of WB-RBS lies in its efficiency, high specificity, and comprehensive anatomical coverage.

This study has several limitations. First, the sample sizes of both the patient and healthy control cohorts were relatively small; validation of reliability requires larger-scale, multi-center studies. Second, not all lesions had histological confirmation. We established the reference standard via consensus based on WB-MRI, which may potentially introduce consensus bias. Third, supplementary imaging may still be needed for osteoblastic metastases, small metastases, and rib metastases. The next phase of research plans to develop an AI-assisted lesion detection module to address this limitation. Fourth, as the WB-RBS sequence was not previously used for diagnostic purposes, establishing clear diagnostic criteria and providing more detailed training for radiologists prior to implementation are crucial for improving diagnostic efficiency. Fifth, reactive hyperplastic hematopoietic marrow due to conditions like anemia or other hematologic disorders can cause heterogeneous bone marrow signal. Although the core positive signs relied upon by this protocol—such as focal nodular cortical destruction, bone defects, and abnormal soft-tissue signal—are less likely to be obscured, further validation with designed subgroups is warranted. Additionally, the healthy volunteer cohort should include more cases with metastasis-mimicking conditions (e.g., osteoporosis, inflammatory bone disease, fractures) to more comprehensively validate the specificity of the diagnostic protocol.

In conclusion, the novel WB-RBS protocol demonstrated good reliability across radiologists of varying experience, different diagnostic confidence thresholds, and patients with lesions of diverse sizes and locations. This protocol provides a feasible, efficient, and readily implementable MRI strategy for bone metastasis screening, significantly reducing examination time and associated costs. Risk-stratifying patients based on WB-RBS screening results may facilitate a more precise and efficient diagnostic pathway. Further optimization through multicenter, large-sample studies, combined with artificial intelligence-assisted analysis, will advance the management of bone metastases into a new era of precise, rapid, and radiation-free whole-body MRI.

## Data availability

The data generated in this study are available upon request from the corresponding author.

## Acknowledgments

We sincerely acknowledge Professor Shaoyu Wang from the MRI Research Collaboration Department of Siemens Healthineers for his guidance on the design of sequence parameters as well as his valuable advice during the manuscript preparation. We also sincerely appreciate all malignant tumor patients and healthy controls who participated in this study; their cooperation made this research possible.

## Abbreviations

AI: Artificial intelligence
BS: Bone scintigraphy
CI: Confidence interval
CT: Computerized tomography
DWI: Diffusion weighted imaging
FOV: Field of view
PACS: Picture archiving and communication system
PET: Positron emission tomography
SD: Standard deviation
SRE: Skeletal-related events
T1WI: T1-weighted image
T2WI: T2-weighted image
TE: Echo Time
TR: Repetition Time
WB-MRI: Whole-body magnetic resonance imaging
WB-RBS: Whole-body rapid bone screening

---

**Supplementary Video S1.** Whole-body coronal images of WB-RBS.

**Supplementary Video S2.** Whole-body sagittal images of WB-RBS.

