## Supplementary material for "Efficacy Validation of a Novel MRI-Based Whole-Body Rapid Bone Scan (WB-RBS) Strategy for Diagnosing Bone Metastases: A Prospective Trial": Supplementary Information.docx

**Content:**

·Methods

·Results

·Supplementary Table S1. Scanning sequence parameters for whole-body MRI, T1-weighted Imaging (T1WI), T2-weighted Imaging (T2WI), and diffusion weighted imaging (DWI).

·Supplementary Table S2. Establishment of four binary classification diagnostic thresholds and binary conversion of scores to metastasis/non-metastasis for diagnostic performance analysis at different confidence levels.

·Supplementary Table S3. Baseline and Follow-up MRI Scanning Status.

·Supplementary Table S4. Inter-reader agreement among three readers for per-patient and per-lesion assessments on WB-MRI and WB-RBS protocols at different thresholds.

·Supplementary Table S5. Agreement between WB-RBS and WB-MRI protocols for per-patient and per-lesion assessments by three readers at different thresholds.

·Supplementary Table S6. Diagnostic performance of three readers for the WB-MRI protocol at patient and lesion levels across different thresholds.

·Supplementary Table S7. Diagnostic performance of the WB-RBS protocol by three readers for lesions of different sizes at different thresholds.

·Supplementary Table S8. Diagnostic performance of the WB-RBS protocol by three readers for lesions in different locations at different thresholds.

·Supplementary Table S9. Correlation of diagnostic results between the gold standard and the three reviewers at four threshold values.

**Methods**

MRI Data Acquisition

The WB-RBS protocol utilized a optimized free breathing FLASH sequence (a fast gradient echo sequence) to acquire images. This included sagittal and coronal views of the skull, thorax and shoulder girdle bones (including sternum, ribs, clavicles, and scapulae), spine (cervical, thoracic, lumbar, and sacrococcygeal vertebrae), pelvis (the bony ring formed by bilateral hip bones and sacrum), and long bones of the limbs (humerus, radius/ulna, femur, tibia/fibula), with additional axial sequences for the chest and pelvis scans to optimize visualization of rib and pelvic bone lesions. The WB-RBS sequence parameters were as follows: repetition time (TR) 7.8 ms, echo time (TE) 3.69 ms, field of view (FOV) 500 × 500 mm², slice thickness 5 mm, voxel size 1 × 1 × 5 mm³, flip angle 20°. Totally, we acquire 5 sagittal scan series, 5 coronal scan series, and 2 axial scan series to ensure full-body bone coverage. After the acquisition of the multi-series sagittal and coronal scans, a large-range full-body mosaic reconstruction is completed via post-processing. Although the theoretical scan time is about 7 minutes, the total scan time is approximately 10 minutes when including shimming and preparation procedures for all sequences before scanning.

The WB-MRI protocol comprised the following sequences: skull (axial T1WI, T2WI), long bones of the limbs (coronal fat-suppressed T1WI and T2WI of the humerus, radius/ulna, femur, tibia/fibula), chest (axial T1WI, T2WI), pelvis (axial fat-suppressed T1WI, T2WI), spine (sagittal fat-suppressed T1WI, T2WI), and axial whole-body diffusion-weighted imaging (DWI) from the head vertex to the distal lower limbs, with a total scan time of approximately 70 minutes. Detailed scan parameters are provided in the Supplementary Table S1.

To enable imaging of the upper limb bones (including humerus, radius, and ulna), a special positioning setup was implemented. For the WB-RBS scan, patients were placed in a “hugging” position with arms crossed and folded against the chest. Despite using a 60-cm bore MRI system, this positioning effectively reduced the required transverse FOV by bringing the arms close to the torso, successfully including the proximal and mid-portions of both upper limbs within the scan coverage. Furthermore, the FLASH sequence employed demonstrated high tolerance to magnetic field inhomogeneity and motion artifacts, ensuring diagnostic-quality bone images could be obtained under this specific positioning and during free breathing. In contrast, the WB-MRI protocol based on T1WI, T2WI, and DWI sequences, being more sensitive to motion and geometric distortion, did not utilize this positioning for scanning.

**Results**

Diagnostic specificity of WB-RBS in healthy individuals

Additionally, the specificity of WB-RBS was validated in 19 healthy individuals without bone metastases. Readers A and B correctly identified all healthy individuals as “no metastasis” at all thresholds. Reader C, however, diagnosed one "metastatic" lesion in each of 2 healthy individuals, but only at the highest threshold (threshold 4), resulting in a false positive rate of approximately 10.5% and a specificity of 89.5%.

**Supplementary Table S1.** Scanning sequence parameters for whole-body MRI, T1-weighted Imaging (T1WI), T2-weighted Imaging (T2WI), and diffusion weighted imaging (DWI).

| **Serial number** | **Part** | **Sequence name** | **Scan orientation** | **TR (ms)** | **TE (ms)** | **FOV (mm^2^)** | **Slice thickness (mm)** |
| --- | --- | --- | --- | --- | --- | --- | --- |
| 1 | Head | T1WI dark-fluid | Axial | 2000 | 19 | 230×230 | 5 |
| 2 |  | T2WI | Axial | 5000 | 95 | 230×230 | 5 |
| 3 | Spine | T1WI | Sagittal | 600 | 9.5 | 450×450 | 4 |
| 4 |  | T2WI fat saturation-dixon | Sagittal | 3270 | 81 | 450×450 | 4 |
| 5 | Thorax and shoulder girdle bones | T1WI | Axial | 3.97 | 1.29 | 380×380 | 4 |
| 6 |  | T2WI fat saturation-fblade trig | Axial | 3535.53 | 80 | 380×380 | 5 |
| 7 | Pelvis | T1WI | Axial | 693 | 12 | 400×400 | 5 |
| 8 |  | T2WI | Axial | 7810 | 81 | 400×400 | 5 |
| 9 | Long bones of the limbs | T1WI | Coronal | 460 | 11 | 500×500 | 5 |
| 10 |  | T2WI fat saturation-dixon | Coronal | 2200 | 62 | 500×500 | 5 |
| 11 | Whole body | DWI (b = 0 and b = 1000 s/mm²) | Axial | 4000 | 54 | 480×480 | 5 |

**Supplementary Table S2.** Establishment of four binary classification diagnostic thresholds and binary conversion of scores to metastasis/non-metastasis for diagnostic performance analysis at different confidence levels.

| **Threshold** | **Score for Metastasis** | **Score for Non-metastasis** |
| --- | --- | --- |
| Thr 1 | 1 | 2，3，4 |
| Thr 2 | 1，2 | 3，4 |
| Thr 3 | 1，2，3 | 4 |
| Thr 4 | 1，2，3，4 | - |

Note: Thr, threshold.

**Supplementary Table S3**. Baseline and follow-up MRI scanning status.

|  | **Number of Cases (n=70)** |
| --- | --- |
| **Baseline scan only** | 53 |
| **Follow-up scan(s)** | 17 |
| **1 follow-up scan** | 17 |
| **2 follow-up scans** | 2 |
| **3 follow-up scans** | 1 |
| **Mean follow-up interval (days)** | 165.1 ± 87.1 |

**Supplementary Table S4**. Inter-reader agreement among three readers for per-patient and per-lesion assessments on WB-MRI and WB-RBS protocols at different thresholds.

|  |  | **WB-MRI** | | **WB-RBS** | |
| --- | --- | --- | --- | --- | --- |
| Readers | Consistency level | Fleiss' κ value | 95%CI | Fleiss' κ value | 95%CI |
| A1，B1，C1 | Patient level | 0.97 | (0.89-1.00) | 0.92 | (0.80-1.00) |
| A2，B2，C2 | Patient level | 1.00 | (1.00-1.00) | 0.90 | (0.79-0.98) |
| A3，B3，C3 | Patient level | 0.97 | (0.90-1.00) | 0.86 | (0.75-0.96) |
| A4，B4，C4 | Patient level | 0.92 | (0.81-1.00) | 0.88 | (0.76-0.96) |
| A1，B1，C1 | Lesion level | 0.95 | (0.89-1.00) | 0.84 | (0.76-0.92) |
| A2，B2，C2 | Lesion level | 1.00 | (1.00-1.00) | 0.84 | (0.75-0.93) |
| A3，B3，C3 | Lesion level | 0.97 | (0.91-1.00) | 0.81 | (0.69-0.91) |
| A4，B4，C4 | Lesion level | 0.93 | (0.85-0.98) | 0.77 | (0.61-0.88) |

Note: The p-values are all less than 0.05. A1-4 indicates the results of Reader A using thresholds 1-4, and B1-4 and C1-4 are similar.

**Supplementary Table S5**. Agreement between WB-RBS and WB-MRI protocols for per-patient and per-lesion assessments by three readers at different thresholds.

| Readers | Consistency level | Cohen' s κ value | Standard error | P value | Consistency level | Cohen' s κ value | Standard error | P value |
| --- | --- | --- | --- | --- | --- | --- | --- | --- |
| A1 | Patient level | 0.83 | 0.08 | <0.001 | Lesion level | 0.58 | 0.08 | <0.001 |
| B1 | Patient level | 0.87 | 0.08 | <0.001 | Lesion level | 0.58 | 0.08 | <0.001 |
| C1 | Patient level | 0.83 | 0.09 | <0.001 | Lesion level | 0.47 | 0.09 | <0.001 |
| A2 | Patient level | 0.84 | 0.08 | <0.001 | Lesion level | 0.53 | 0.09 | <0.001 |
| B2 | Patient level | 0.80 | 0.09 | <0.001 | Lesion level | 0.45 | 0.10 | <0.001 |
| C2 | Patient level | 0.76 | 0.10 | <0.001 | Lesion level | 0.36 | 0.10 | <0.001 |
| A3 | Patient level | 0.77 | 0.09 | <0.001 | Lesion level | 0.35 | 0.10 | <0.001 |
| B3 | Patient level | 0.67 | 0.10 | <0.001 | Lesion level | 0.28 | 0.10 | 0.006 |
| C3 | Patient level | 0.70 | 0.10 | <0.001 | Lesion level | 0.25 | 0.10 | 0.014 |
| A4 | Patient level | 0.61 | 0.10 | <0.001 | Lesion level | 0.16 | 0.10 | 0.098 |
| B4 | Patient level | 0.55 | 0.10 | <0.001 | Lesion level | 0.00 | 0.09 | 0.974 |
| C4 | Patient level | 0.57 | 0.10 | <0.001 | Lesion level | 0.02 | 0.09 | 0.827 |

Note: The p-values are all less than 0.05. A1-4 indicates the results of Reader A using thresholds 1-4, and B1-4 and C1-4 are similar.

**Supplementary Table S6**. Diagnostic performance of three readers for the WB-MRI protocol at patient and lesion levels across different thresholds.

| Readers | Level | Sensitivity (%) | Specificity (%) | PPV (%) | NPV (%) | Accuracy (%) |
| --- | --- | --- | --- | --- | --- | --- |
| A1 | Patient level | 93.3 | 98.2 | 93.3 | 98.2 | 97.1 |
| A2 | Patient level | 93.3 | 98.2 | 93.3 | 98.2 | 97.1 |
| A3 | Patient level | 100.0 | 98.2 | 93.8 | 100.0 | 98.6 |
| A4 | Patient level | 100.0 | 98.2 | 93.8 | 100.0 | 98.6 |
| B1 | Patient level | 86.7 | 98.2 | 92.9 | 96.4 | 95.7 |
| B2 | Patient level | 93.3 | 98.2 | 93.3 | 98.2 | 97.1 |
| B3 | Patient level | 93.3 | 98.2 | 93.3 | 98.2 | 97.1 |
| B4 | Patient level | 100.0 | 96.4 | 88.2 | 100.0 | 97.1 |
| C1 | Patient level | 86.7 | 98.2 | 92.9 | 96.4 | 95.7 |
| C2 | Patient level | 93.3 | 98.2 | 93.3 | 98.2 | 97.1 |
| C3 | Patient level | 93.3 | 98.2 | 93.3 | 98.2 | 97.1 |
| C4 | Patient level | 93.3 | 94.5 | 82.4 | 98.1 | 94.3 |
| A1 | Lesion level | 98.0 | 100.0 | 100.0 | 92.6 | 98.4 |
| A2 | Lesion level | 99.0 | 100.0 | 100.0 | 96.2 | 99.2 |
| A3 | Lesion level | 100.0 | 100.0 | 100.0 | 100.0 | 100.0 |
| A4 | Lesion level | 100.0 | 100.0 | 100.0 | 100.0 | 100.0 |
| B1 | Lesion level | 96.0 | 100.0 | 100.0 | 86.2 | 96.8 |
| B2 | Lesion level | 99.0 | 100.0 | 100.0 | 96.2 | 99.2 |
| B3 | Lesion level | 99.0 | 100.0 | 100.0 | 96.2 | 99.2 |
| B4 | Lesion level | 100.0 | 96.0 | 99.0 | 100.0 | 99.2 |
| C1 | Lesion level | 96.0 | 100.0 | 100.0 | 86.2 | 96.8 |
| C2 | Lesion level | 99.0 | 100.0 | 100.0 | 96.2 | 99.2 |
| C3 | Lesion level | 99.0 | 96.0 | 99.0 | 96.0 | 98.4 |
| C4 | Lesion level | 99.0 | 88.0 | 97.1 | 95.7 | 96.8 |

Note: A1-4 indicates the results of Reader A using thresholds 1-4, and B1-4 and C1-4 are similar.

**Supplementary Table S7.** Diagnostic performance of the WB-RBS protocol by three readers for lesions of different sizes at different thresholds.

| **Lesion Size** | **Readers** | **Sensitivity (%)** | **Specificity (%)** | **PPV (%)** | **NPV (%)** | **Accuracy (%)** |
| --- | --- | --- | --- | --- | --- | --- |
| ≤1.5cm | A1 | 72.1 | 88.9 | 95.7 | 48.5 | 75.9 |
| ≤1.5cm | A2 | 80.3 | 66.7 | 89.1 | 50.0 | 77.2 |
| ≤1.5cm | A3 | 82.0 | 50.0 | 84.7 | 45.0 | 74.7 |
| ≤1.5cm | A4 | 82.0 | 27.8 | 79.4 | 31.2 | 69.6 |
| ≤1.5cm | B1 | 67.2 | 88.9 | 95.3 | 44.4 | 72.2 |
| ≤1.5cm | B2 | 78.7 | 66.7 | 88.9 | 48.0 | 75.9 |
| ≤1.5cm | B3 | 82.0 | 38.9 | 82.0 | 38.9 | 72.2 |
| ≤1.5cm | B4 | 82.0 | 16.7 | 76.9 | 21.4 | 67.1 |
| ≤1.5cm | C1 | 65.6 | 77.8 | 90.9 | 40.0 | 68.4 |
| ≤1.5cm | C2 | 72.1 | 61.1 | 86.3 | 39.3 | 69.6 |
| ≤1.5cm | C3 | 77.0 | 50.0 | 83.9 | 39.1 | 70.9 |
| ≤1.5cm | C4 | 77.0 | 22.2 | 77.0 | 22.2 | 64.6 |
| >1.5cm | A1 | 95.0 | 71.4 | 95.0 | 71.4 | 91.5 |
| >1.5cm | A2 | 95.0 | 71.4 | 95.0 | 71.4 | 91.5 |
| >1.5cm | A3 | 95.0 | 42.9 | 90.5 | 60.0 | 87.2 |
| >1.5cm | A4 | 95.0 | 28.6 | 88.4 | 50.0 | 85.1 |
| >1.5cm | B1 | 95.0 | 71.4 | 95.0 | 71.4 | 91.5 |
| >1.5cm | B2 | 95.0 | 42.9 | 90.5 | 60.0 | 87.2 |
| >1.5cm | B3 | 95.0 | 28.6 | 88.4 | 50.0 | 85.1 |
| >1.5cm | B4 | 95.0 | 0.0 | 84.4 | 0.0 | 80.9 |
| >1.5cm | C1 | 95.0 | 42.9 | 90.5 | 60.0 | 87.2 |
| >1.5cm | C2 | 95.0 | 42.9 | 90.5 | 60.0 | 87.2 |
| >1.5cm | C3 | 95.0 | 0.0 | 84.4 | 0.0 | 80.9 |
| >1.5cm | C4 | 95.0 | 0.0 | 84.4 | 0.0 | 80.9 |

Note: A1-4 indicates the results of Reader A using thresholds 1-4, and B1-4 and C1-4 are similar. PPV, positive predictive value; NPV, negative predictive value.

**Supplementary Table S8.** Diagnostic performance of the WB-RBS protocol by three readers for lesions in different locations at different thresholds.

| **Lesion Location** | **Readers** | **Sensitivity (%)** | **Specificity (%)** | **PPV (%)** | **NPV (%)** | **Accuracy (%)** |
| --- | --- | --- | --- | --- | --- | --- |
| Thoracic cage and shoulder | A1 | 45.5 | 0.0 | 100.0 | 0.0 | 45.5 |
| Thoracic cage and shoulder | A2 | 45.5 | 0.0 | 100.0 | 0.0 | 45.5 |
| Thoracic cage and shoulder | A3 | 45.5 | 0.0 | 100.0 | 0.0 | 45.5 |
| Thoracic cage and shoulder | A4 | 45.5 | 0.0 | 100.0 | 0.0 | 45.5 |
| Thoracic cage and shoulder | B1 | 45.5 | 0.0 | 100.0 | 0.0 | 45.5 |
| Thoracic cage and shoulder | B2 | 45.5 | 0.0 | 100.0 | 0.0 | 45.5 |
| Thoracic cage and shoulder | B3 | 45.5 | 0.0 | 100.0 | 0.0 | 45.5 |
| Thoracic cage and shoulder | B4 | 45.5 | 0.0 | 100.0 | 0.0 | 45.5 |
| Thoracic cage and shoulder | C1 | 36.4 | 0.0 | 100.0 | 0.0 | 36.4 |
| Thoracic cage and shoulder | C2 | 36.4 | 0.0 | 100.0 | 0.0 | 36.4 |
| Thoracic cage and shoulder | C3 | 36.4 | 0.0 | 100.0 | 0.0 | 36.4 |
| Thoracic cage and shoulder | C4 | 36.4 | 0.0 | 100.0 | 0.0 | 36.4 |
| Spine | A1 | 78.1 | 100.0 | 100.0 | 68.2 | 85.1 |
| Spine | A2 | 90.6 | 80.0 | 90.6 | 80.0 | 87.2 |
| Spine | A3 | 90.6 | 46.7 | 78.4 | 70.0 | 76.6 |
| Spine | A4 | 90.6 | 26.7 | 72.5 | 57.1 | 70.2 |
| Spine | B1 | 81.2 | 100.0 | 100.0 | 71.4 | 87.2 |
| Spine | B2 | 90.6 | 66.7 | 85.3 | 76.9 | 83.0 |
| Spine | B3 | 90.6 | 33.3 | 74.4 | 62.5 | 72.3 |
| Spine | B4 | 90.6 | 13.3 | 69.0 | 40.0 | 66.0 |
| Spine | C1 | 78.1 | 66.7 | 83.3 | 58.8 | 74.5 |
| Spine | C2 | 87.5 | 53.3 | 80.0 | 66.7 | 76.6 |
| Spine | C3 | 90.6 | 26.7 | 72.5 | 57.1 | 70.2 |
| Spine | C4 | 90.6 | 6.7 | 67.4 | 25.0 | 63.8 |
| Pelvis | A1 | 91.2 | 55.6 | 88.6 | 62.5 | 83.7 |
| Pelvis | A2 | 91.2 | 44.4 | 86.1 | 57.1 | 81.4 |
| Pelvis | A3 | 91.2 | 44.4 | 86.1 | 57.1 | 81.4 |
| Pelvis | A4 | 91.2 | 33.3 | 83.8 | 50.0 | 79.1 |
| Pelvis | B1 | 82.4 | 55.6 | 87.5 | 45.5 | 76.7 |
| Pelvis | B2 | 88.2 | 44.4 | 85.7 | 50.0 | 79.1 |
| Pelvis | B3 | 91.2 | 33.3 | 83.8 | 50.0 | 79.1 |
| Pelvis | B4 | 91.2 | 11.1 | 79.5 | 25.0 | 74.4 |
| Pelvis | C1 | 82.4 | 66.7 | 90.3 | 50.0 | 79.1 |
| Pelvis | C2 | 85.3 | 55.6 | 87.9 | 50.0 | 79.1 |
| Pelvis | C3 | 85.3 | 44.4 | 85.3 | 44.4 | 76.7 |
| Pelvis | C4 | 85.3 | 33.3 | 82.9 | 37.5 | 74.4 |
| Long bones of limbs | A1 | 87.5 | 100.0 | 100.0 | 25.0 | 88.0 |
| Long bones of limbs | A2 | 91.7 | 100.0 | 100.0 | 33.3 | 92.0 |
| Long bones of limbs | A3 | 95.8 | 100.0 | 100.0 | 50.0 | 96.0 |
| Long bones of limbs | A4 | 95.8 | 0.0 | 95.8 | 0.0 | 92.0 |
| Long bones of limbs | B1 | 83.3 | 100.0 | 100.0 | 20.0 | 84.0 |
| Long bones of limbs | B2 | 91.7 | 100.0 | 100.0 | 33.3 | 92.0 |
| Long bones of limbs | B3 | 95.8 | 100.0 | 100.0 | 50.0 | 96.0 |
| Long bones of limbs | B4 | 95.8 | 0.0 | 95.8 | 0.0 | 92.0 |
| Long bones of limbs | C1 | 87.5 | 100.0 | 100.0 | 25.0 | 88.0 |
| Long bones of limbs | C2 | 87.5 | 100.0 | 100.0 | 25.0 | 88.0 |
| Long bones of limbs | C3 | 95.8 | 100.0 | 100.0 | 50.0 | 96.0 |
| Long bones of limbs | C4 | 95.8 | 0.0 | 95.8 | 0.0 | 92.0 |

Note: A1-4 indicates the results of Reader A using thresholds 1-4, and B1-4 and C1-4 are similar. PPV, positive predictive value; NPV, negative predictive value.

**Supplementary Table 9.** Correlation of diagnostic results between the gold standard and the three reviewers at four threshold values.

| **Variable Pairs** | **Level** | **Correlation Coefficient** | **P value** |
| --- | --- | --- | --- |
| Gold Standard & A1 | Patient level | 0.75 | < 0.001 |
| Gold Standard & A2 | Patient level | 0.76 | < 0.001 |
| Gold Standard & A3 | Patient level | 0.73 | < 0.001 |
| Gold Standard & A4 | Patient level | 0.60 | < 0.001 |
| Gold Standard & B1 | Patient level | 0.75 | < 0.001 |
| Gold Standard & B2 | Patient level | 0.73 | < 0.001 |
| Gold Standard & B3 | Patient level | 0.62 | < 0.001 |
| Gold Standard & B4 | Patient level | 0.52 | < 0.001 |
| Gold Standard & C1 | Patient level | 0.71 | < 0.001 |
| Gold Standard & C2 | Patient level | 0.68 | < 0.001 |
| Gold Standard & C3 | Patient level | 0.65 | < 0.001 |
| Gold Standard & C4 | Patient level | 0.54 | < 0.001 |
| Gold Standard & A1 | Lesion level | 0.56 | < 0.001 |
| Gold Standard & A2 | Lesion level | 0.50 | < 0.001 |
| Gold Standard & A3 | Lesion level | 0.35 | < 0.001 |
| Gold Standard & A4 | Lesion level | 0.17 | 0.065 |
| Gold Standard & B1 | Lesion level | 0.52 | < 0.001 |
| Gold Standard & B2 | Lesion level | 0.42 | < 0.001 |
| Gold Standard & B3 | Lesion level | 0.24 | 0.006 |
| Gold Standard & B4 | Lesion level | -0.01 | 0.908 |
| Gold Standard & C1 | Lesion level | 0.39 | < 0.001 |
| Gold Standard & C2 | Lesion level | 0.34 | < 0.001 |
| Gold Standard & C3 | Lesion level | 0.20 | 0.024 |
| Gold Standard & C4 | Lesion level | 0.00 | 0.985 |
